# Frequency of Dengue Virus-Specific T Cells is related to Infection Outcome in Endemic Settings

**DOI:** 10.1101/2024.02.05.24302330

**Authors:** Rosa Isela Gálvez, Amparo Martínez-Pérez, E. Alexandar Escarrega, Tulika Singh, José Víctor Zambrana, Ángel Balmaseda, Eva Harris, Daniela Weiskopf

**Affiliations:** Center for Vaccine Innovation, La Jolla Institute for Immunology, La Jolla, CA 92037, USA; Division of Infectious Diseases and Vaccinology, School of Public Health, University of California, Berkeley, Berkeley, CA 94720-3370, USA; Sustainable Sciences Institute, Managua, Nicaragua; Department of Epidemiology, School of Public Health, University of Michigan, Ann Arbor, MI 48109-2029, USA; Laboratorio Nacional de Virología, Centro Nacional de Diagnóstico y Referencia, Ministerio de Salud, Managua, Nicaragua; Division of Infectious Diseases and Global Public Health, School of Medicine, University of California, San Diego, La Jolla, CA 92037, USA

## Abstract

Dengue is widespread in tropical and subtropical regions globally and leads to a considerable burden of disease. Annually, dengue virus (DENV) causes up to 400 million infections, of which ~25% present with clinical symptoms ranging from mild to fatal. Despite its significance as a growing public health concern, the development of effective DENV vaccines has been highly challenging. One of the reasons is the lack of comprehensive understanding of the influence exerted by prior DENV infections and immune responses with cross-reactive properties. To investigate this, we collected samples from a pediatric cohort study in dengue-endemic Managua, Nicaragua. We characterized T cell responses in a group of 71 healthy children who had previously experienced one or more natural DENV infections and who, within one year after sample collection, had a subsequent DENV infection that was either symptomatic (n=25) or inapparent (n=46, absence of clinical disease). Thus, our study was designed to investigate the impact of pre-existing DENV specific T cell responses on the clinical outcomes of subsequent DENV infection. We assessed the DENV specific T cell responses using an activation-induced marker assay (AIM). Children who had experienced only one prior DENV infection displayed heterogeneous DENV specific CD4^+^ and CD8^+^ T cell frequencies. In contrast, children who had experienced two or more DENV infections showed significantly higher frequencies of DENV specific CD4^+^ and CD8^+^ T cells that were associated with inapparent as opposed to symptomatic outcomes in the subsequent DENV infection. Taken together, these findings demonstrate the protective role of DENV specific T cells against symptomatic DENV infection and constitute an advancement toward identifying protective immune correlates against dengue fever and clinical disease.

**Graphical Abstract:** 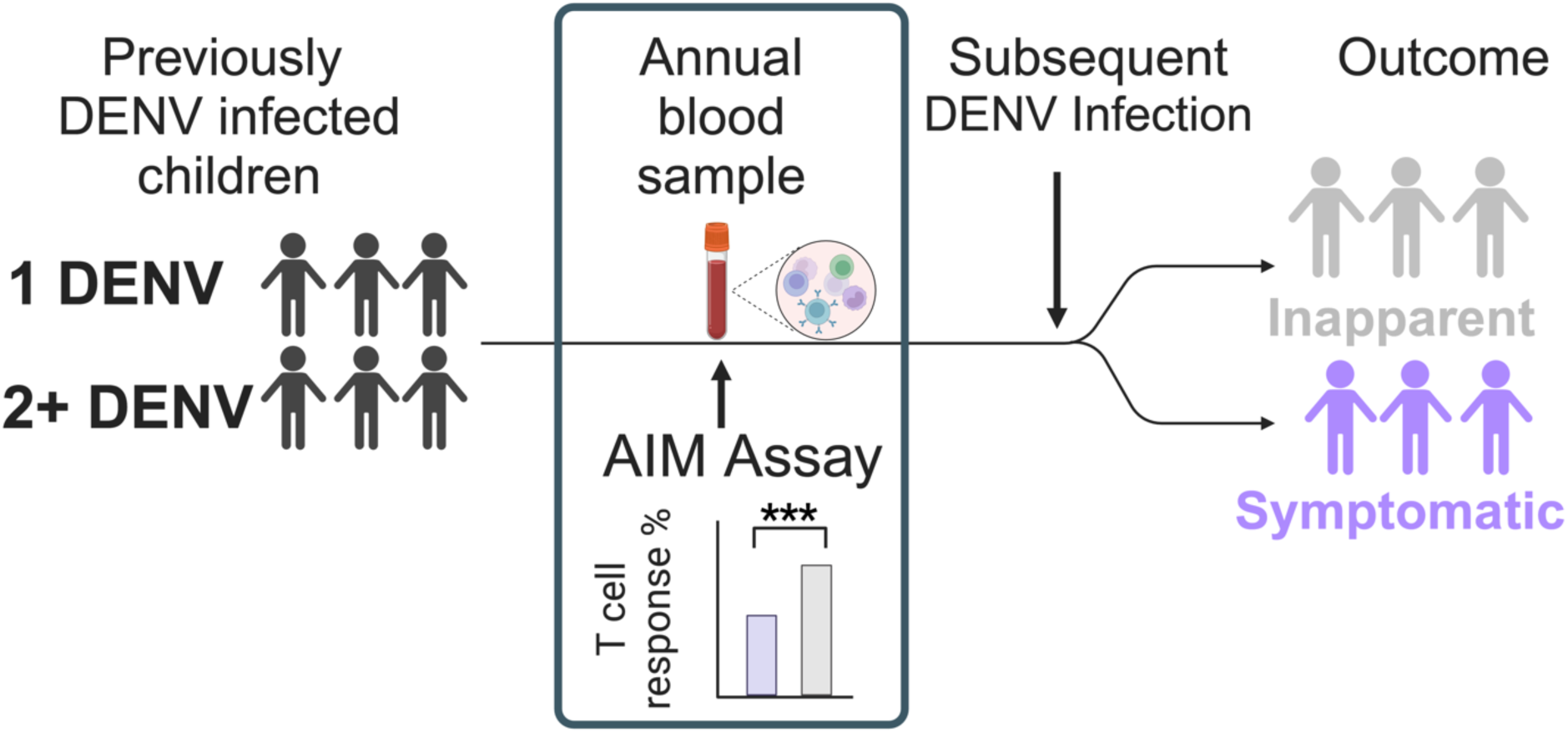

## Introduction

In 2023, the WHO declared dengue to be the world’s fastest-spreading tropical disease and to represent a “pandemic threat” to nearly half of the global population [1]. Dengue is caused by four dengue virus (DENV) serotypes, which are enveloped viruses containing a single-stranded, positive-sense RNA genome [2]. Dengue viruses circulate worldwide in the tropics and subtropics and annually cause an estimated 400 million DENV infections, with up to 100 million resulting in clinical cases and at least 22,000 deaths [3]. Understanding the sequential development of immunity to dengue viruses is crucial since individuals living in endemic regions are sequentially exposed to multiple serotypes. The precise immunopathogenic mechanisms related to sequential DENV infections with a different serotype (heterologous infection) remain only partially elucidated [4]. Substantial evidence suggests that the humoral arm of the adaptive immune response, triggered by the initial infection, plays a role in both protecting against disease and promoting infection, and enhancing DENV replication during subsequent infection [5–7]. Cohort studies have demonstrated that in cases of secondary heterologous DENV infection, the duration of viremia and, more importantly, infection and disease outcome are influenced by the level of pre-existing antibodies against DENV such that a specific range of antibody titers is associated with the likelihood of protection or the risk of experiencing severe dengue during a subsequent infection [8]. Interestingly, this relation appears to depend on the serotype of the subsequent infection [7, 9]. The underlying distinct mechanisms are still yet to be investigated.

In a similar manner to the humoral response, the adaptive cellular responses exerted by T cells are acknowledged to play a pivotal role in the control of DENV infections [4, 10, 11]. T cells provide durable protective immunity [12], yet they have also been linked to eliciting immune-mediated damage [13]. However, the precise factors governing the establishment of memory T cell responses in the context of multiple DENV infections are still in need of comprehensive elucidation. The presence of pre-existing T cell memory has the potential to significantly influence outcomes during secondary heterologous DENV infections, promoting faster and more robust T cell responses as compared to those encountered during primary infection [14]. However, these features of T cells have also been speculated to elevate the susceptibility of individuals to severe forms of dengue fever and clinical illness [15, 16].

In this context, the goal of our study was to assess the relationship between the magnitude of naturally acquired T cell memory resulting from one or more DENV infections and their impact on a subsequent DENV infection. This study aimed to expand our understanding of the role of pre-existing DENV specific T cells in either providing protective immunity or triggering immunopathogenic responses leading to clinical disease. We hypothesized that a higher pre-existing frequency of CD4^+^ and CD8^+^ T cells would be associated with protection from subsequent symptomatic DENV infection. We analyzed T cell responses using flow cytometry, evaluating the phenotypes and frequencies of CD4^+^ and CD8^+^ T cells individually, and examining relationships with the outcome of subsequent DENV infection. Importantly, we were able to test our question using a substantial sample size (n=71), together with accurate characterization of infection history, information about the HLA type of the donors, and our established DENV peptide MegaPool approach (MP) to asses DENV specific T cell responses [17]. We hereby provide a comprehensive assessment of T cell memory in multiple DENV infections in an endemic setting.

## Results

### Pediatric cohort characteristics

Our study used pre-infection samples obtained from 71 children enrolled in the Pediatric Dengue Cohort Study, which has been ongoing in Managua, Nicaragua, since 2004. These samples were collected between 2006 and 2019 and grouped based on the outcome of the subsequent DENV infection: 46 participants who subsequently developed inapparent DENV infections versus 25 participants who later experienced symptomatic DENV infection. The mean age was 9 years in the pre-inapparent group and 11 years in the pre-symptomatic group. In the pre-inapparent infection group, 48% of participants were male and 52% were female. In the pre-symptomatic infection group, 64% were male and 36% were female. The age distribution was examined for balance between groups, with a Mann-Whitney test yielding a p-value of 0.0799. Additionally, the association between sex and the pre-inapparent or pre-symptomatic group was assessed using the Chi-square test with Yates correction, and the results indicated no statistically significant difference. In summary, the groups can be characterized as being well-balanced. Furthermore, to categorize the participants, we considered their infection history, distinguishing between those who had encountered only one prior DENV infection (referred to as 1 DENV) and those who had experienced two or more previous DENV infections (referred to as 2+ DENV). Among the individuals in the pre-inapparent infection group (who remained protected against subsequent dengue symptoms), 61% had a history of one previous DENV infection (1 DENV) and 39% had experienced two or more previous infections (2+ DENV). Within the pre-symptomatic group, 44% had experienced one previous DENV infection (1 DENV), while 56% had experienced two or more DENV infections (2+ DENV) **(Table 1)**.

**Table 1.** Demographic participants characteristics.

|  | Pre-Inapparent | Pre-Symptomatic | p |
| --- | --- | --- | --- |
| **Donors, n (%)** | 46 (64.8%) | 25 (35.2%) |  |
| **Age** (years (mean, SD)) | 9, 3.1 | 11, 2.8 | 0.0799 |
| **Sex, n (%)** |  |  | 0.1919 |
| Male | 22 (48%) | 16 (64%) |  |
| Female | 24 (52%) | 9 (36%) |  |
| **Infection history, n (%)** |  |  |  |
| 1 DENV | 28 (61%) | 11(44%) |  |
| 2+ DENV | 18 (39%) | 14 (56%) |  |
| **Years since last infection**<br>(Median + IQR) |  |  |  |
| all samples together<br>(regardless of the # of<br>infections) | 2, IQR 2 | 4, IQR 2 |  |
| 1 DENV | 2, IQR 1.5 | 4, IQR 5 |  |
| 2+ DENV | 4, IQR 2 | 4, IQR 2 |  |

### DENV specific CD4^+^ T cell memory elicited by previous natural DENV infections

DENV specific CD4^+^ T cell memory responses were evaluated following a 24h stimulation of peripheral blood mononuclear cells (PBMCs) with DENV CD4 megapools (MP) and using the Activation Induced Marker Assay (AIM Assay) as readout. Our laboratory has developed the MP approach to study the breadth of T cell responses to diverse pathogens [17]. In previous studies, HLA class I and HLA class II-restricted DENV epitopes spanning the entire sequence of the DENV viruses were predicted and subsequently experimentally tested in different human cohorts from endemic regions [10, 18, 19]. In this study, we have used two different DENV specific MPs, one tailored to measure CD4^+^ T cell responses (containing 180 epitopes) and one tailored to CD8^+^ T cell responses (containing 268 epitopes).

Consequently, the CD4^+^ T cell reactivity to DENV MPs assessed by the AIM assay is represented as upregulation of the markers OX40^+^CD137^+^ on CD4^+^ T cells following activation [20–22]. T cell responses were calculated by subtracting individual CD4^+^ T cell responses to the negative control (PBMCs only incubated with the DMSO solvent for 24h) from each individual’s CD4^+^ T cell response to DENV MP. Upon direct comparison of the geometric mean of the AIM^+^ CD4^+^ T cells between the pre-inapparent and pre-symptomatic group, no statistically significant differences could be detected.

In this study, the term responder is used to describe T cells that exhibit a measurable upregulation of activation markers in response to the DENV MP stimulation. Within the pre-symptomatic group, there was a lower frequency of responders (52%) compared to the pre-inapparent group (67%) **(Figure 1A)**. Next, we stratified groups based on the number of prior DENV infections they had experienced in the past. The frequency of responders was higher in both groups after 2+ DENV infections compared to the frequency of responders after only 1 DENV infection (pre-symptomatic: 45% vs. 57%, and pre-inapparent: 53% vs. 88%). There was no significant difference between the pre-symptomatic and pre-inapparent groups linked to the outcome of subsequent infections following a single DENV exposure. When examining the magnitude of AIM^+^ responses within the pre-inapparent group, a significant difference emerged between those with 1 DENV prior infection and those with 2+ DENV prior infections, suggesting that more than one DENV infection may be necessary to induce a robust CD4^+^ T cell memory response **(Figure 1B)**.

**Figure 1.**
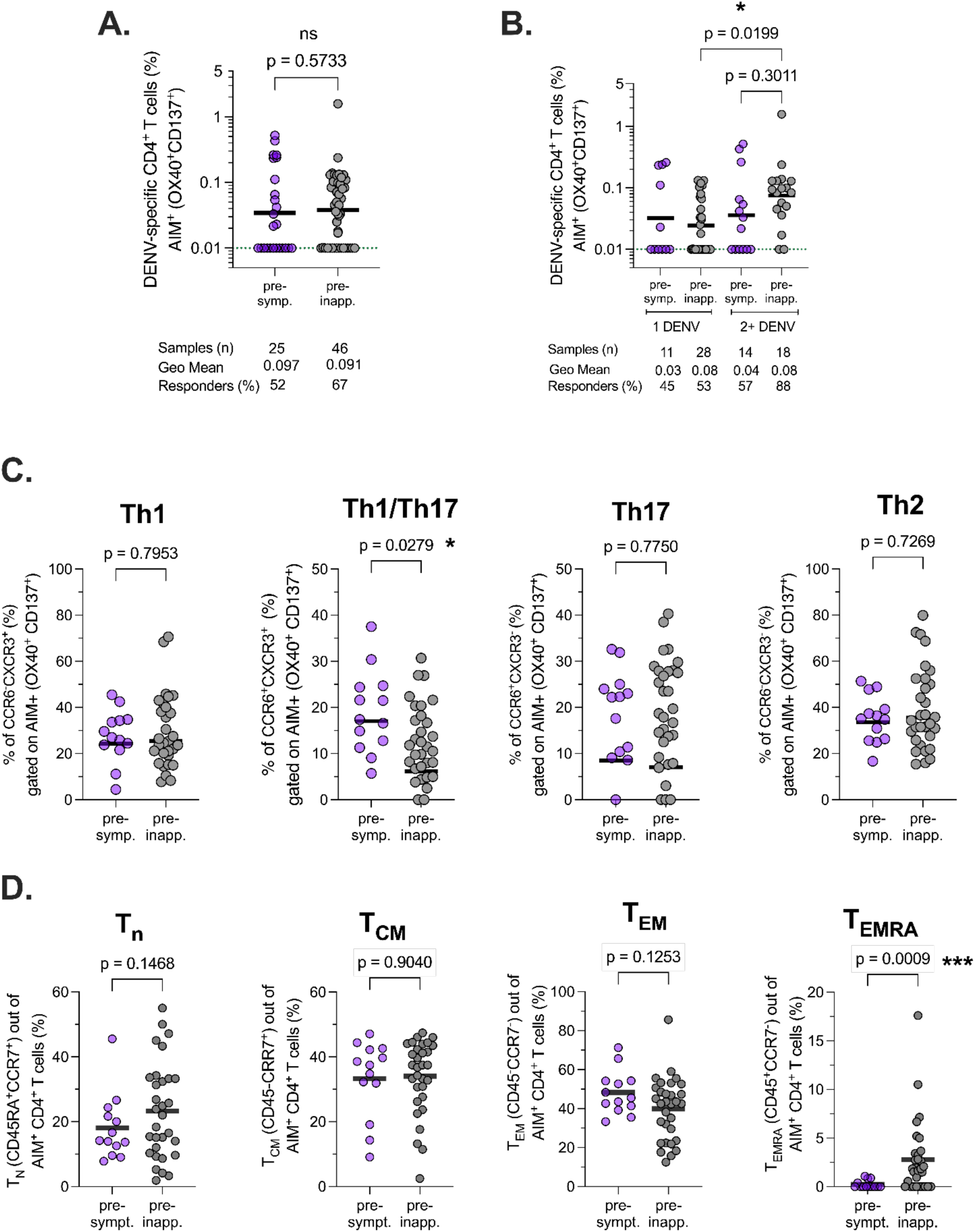
DENV specific CD4^+^ T cell memory elicited by previous natural infections. (**A**) DENV specific CD4^+^ T cell memory responses between pre-inapparent (grey circles) and pre-symptomatic (purple circles) groups were compared. (**B**) DENV specific CD4^+^ T cell responses were stratified based on the number of prior DENV infections. The dotted green line indicates the limit of quantification (LOQ). Baseline and non-responders were set at 0.01 of LOQ. Bars represent the geometric mean. Data were analyzed for statistical significance using the Mann-Whitney test. (**C**) Th cell subset frequency within AIM^+^ CD4^+^ T cells was analyzed based on chemokine receptor expression using CXCR3 and CCR6 as markers. (**D**) Distribution of memory T cell subsets was defined within AIM^+^ CD4^+^ T cells based on CCR7 and CD45RA expression as T cell naive (T_N_), T central memory (T_CM_), T effector memory (T_eff_), and T effector memory re-expressing CD45RA (T_EMRA_). For ancestral gating and representative flow cytometry plots of DENV specific CD4^+^ T cells (OX40^+^CD137^+^), see Supplemental Figure 2.

We further analyzed the phenotype of DENV specific CD4^+^ T cells (AIM^+^) using antibodies against chemokine receptors with preferential expression on functionally distinct memory T cell subsets. This analysis describes four distinct T helper cell subsets (Th) based on chemokine receptor expression: CXCR3^+^CCR6^−^ and CXCR3^+^CCR6^+^ (both enriched in Th1/Th17 cells); CXCR3^−^CCR6^+^ (enriched in Th17 cells) and CXCR3^−^CCR6^−^(enriched in Th2 cells). The pre-inapparent group demonstrated significantly higher levels of the DENV specific memory T cells characterized by CXCR3+CCR6+ expression as compared to the pre-symptomatic group **(Figure 1C)**.

Next, we enumerated the DENV specific CD4^+^ T cell memory subsets by employing a combination of CD45RA and CCR7 markers. We found that in all analyzed participants regardless of the stratification by infection history, the DENV specific CD4^+^ T cells were predominantly composed of effector memory T cells (T_EM_: CD45RA^−^CCR7^−^), followed by central memory T cells (T_CM_: CD45RA^−^CCR7^+^), and then the T effector memory recently activated (T_EMRA_) subset (CCR7^+^CD45RA^+^), which was the smallest subset. Regarding the distribution of these memory T cell subsets, the geometric mean frequency varied. For effector memory T cells, the mean was 47% in the pre-symptomatic group compared to 37% in the pre-inapparent group. Central memory T cells exhibited 30% in the pre-symptomatic group and 31% in the pre-inapparent group, with no significant differences observed between the two groups. However, the T_EMRA_ subset (CCR7^+^CD45RA^+^) was significantly different between the pre-inapparent and pre-symptomatic groups (p=0.009, analyzed with the Mann-Whitney test), constituting a very low percentage (0.25%) in the pre-symptomatic group compared to a significantly higher percentage (2.8%, p=0.0009) in the pre-inapparent group **(Figure 1D)**.

### DENV specific CD8^+^ T cell memory elicited by previous natural DENV infections

Analysis of the memory DENV specific CD8^+^ T cell response was performed after a 24h stimulation with DENV specific CD8 MP and using the AIM assay as readout. After activation, the reactivity of CD8^+^ T cells to DENV MPs, evaluated through the AIM assay, is indicated by an increase in the co-expression of CD69^+^ and CD137^+^ markers on CD8^+^ T cells. Upon direct comparison of the pre-inapparent (grey circles) and pre-symptomatic (purple circles) groups, there were no statistically significant differences between the geometric mean of the AIM^+^ responses: 54% of responders were observed in the pre-symptomatic group and 63% of responders were observed in the pre-inapparent group **(Figure 2A)**.

**Figure 2.**
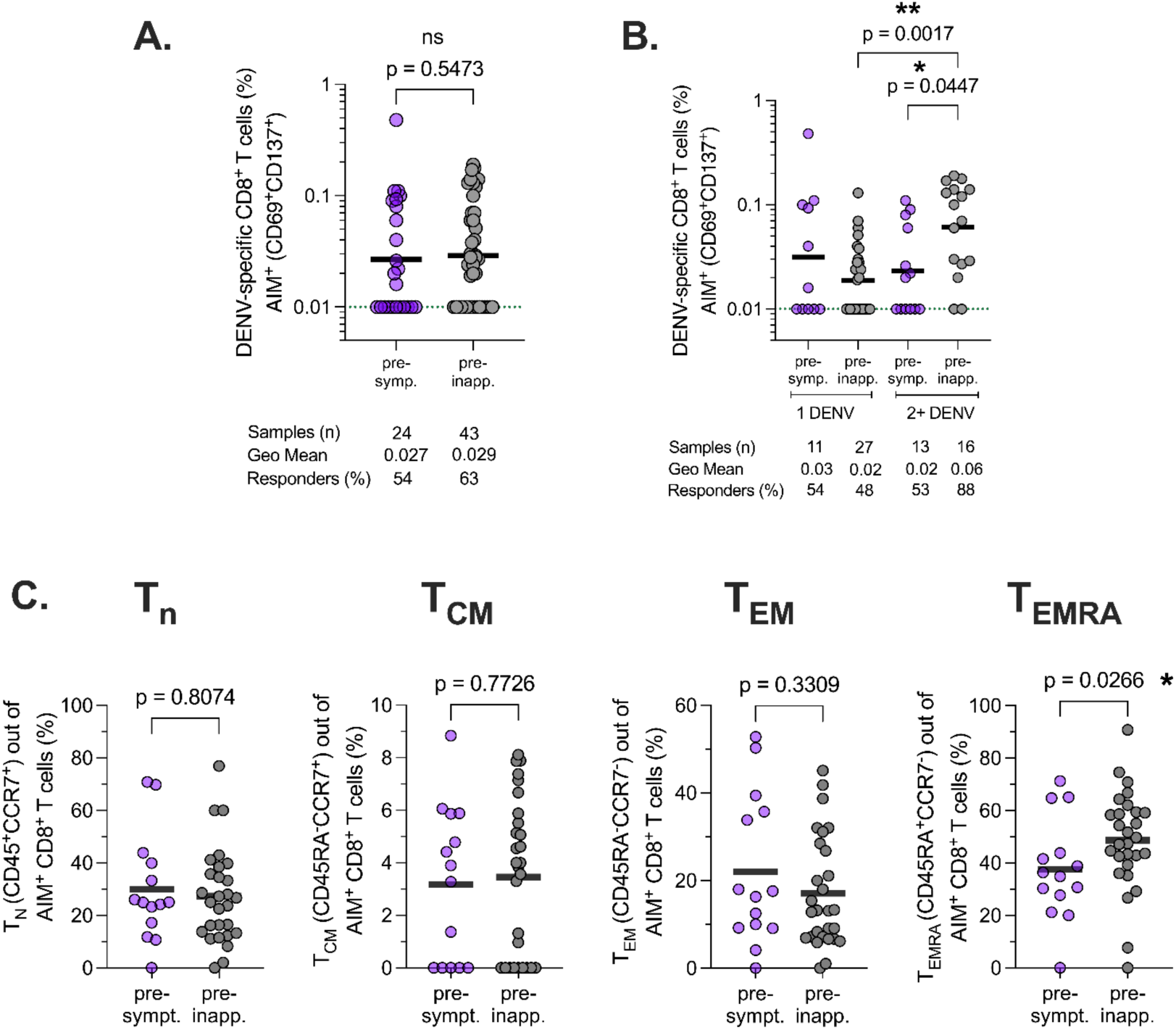
DENV specific CD8^+^ T cell memory elicited by previous natural infections. DENV specific CD8^+^ T cell memory responses were assessed with the AIM assay following a 24h stimulation of PBMCs with DENV CD8 MP. T cell responses were calculated as background-subtracted responses to DENV CD8 MP per individual. The dotted green line indicates the limit of quantification (LOQ). Baseline and non-responders are set at 0.01 of LOQ. Bars represent geometric mean. Data were analyzed for statistical significance using the Mann-Whitney test. (**A**) DENV specific CD8^+^ T cell memory responses between pre-inapparent and pre-symptomatic groups were compared. (**B**) DENV specific CD8^+^ T cell responses were stratified based on the number of prior DENV infections. (**C**) The distribution of memory T cell subsets was defined within AIM^+^ CD4^+^ T cells based on CCR7 and CD45RA expression. For ancestral gating and representative flow cytometry plots of DENV specific CD8^+^ T cells (CD69^+^CD137^+^), see Supplemental Figure 2.

Subsequently, upon stratification of these groups based on the participants’ history of prior DENV infections, differences in the magnitude of the AIM^+^ response once again became apparent. A noteworthy increase in AIM^+^ response within the pre-inapparent group was observed following 2+ DENV infections as compared to the pre-symptomatic group (p=0.0447). Also, in the pre-inapparent group, the frequency of CD8^+^ T cell memory responders was substantially higher after 2+ DENV infections (88%) as compared to only 1 DENV infection (48%). When comparing the magnitude of the AIM^+^ response exclusively within the pre-inapparent group, analogous to the observations made for CD4^+^ T cells, a significant difference between participants who had experienced only 1 DENV infection and those who had encountered 2+ DENV infections was observed (p=0.0017). In conclusion, similar to the findings for DENV specific CD4^+^ T cells, our results suggest that multiple DENV infections may be necessary to trigger a robust DENV specific CD8^+^ T cell memory response **(Figure 2B)**.

Lastly, we investigated the memory subsets of DENV specific CD8^+^ T cells by employing a combination of CD45RA and CCR7 markers. Irrespective of the participants’ infection history, DENV specific CD8^+^ T cells were predominantly composed of the T_EMRA_ subset (CCR7^+^CD45RA^+^). Specifically, this subset constituted 38% of the pre-symptomatic group compared to a higher percentage of 49% in the pre-inapparent group. Additionally, a significant difference was observed within the T_EMRA_ subset (CCR7^+^CD45RA+) when comparing the two groups (p=0.0266, analyzed utilizing the Mann-Whitney test). No significant differences by infection history were observed among the percentages of naïve, central, and effector memory T cell subsets in terms of the geometric mean **(Figure 2C)**.

### Association of AIM^+^ CD4^+^ T cell responses with pre-existing DENV antibody titers

CD4^+^ T cells promote B cell stimulation and consequently B cell-derived antibody responses; thus, we sought to delineate the relationship between pre-existing CD4^+^ T cells and DENV binding antibody titer. We examined the association of the magnitude of AIM^+^ CD4^+^ T cells in pre-symptomatic and pre-inapparent groups with their corresponding pre-existing DENV antibody titers. When comparing the overall magnitude of DENV specific AIM^+^ CD4^+^ T cell responses within bins of DENV iELISA antibody titer, we did not note any statistically significant differences. However, differences emerged when we compared the frequencies of responders (%) in each group. These are visually represented in the lower panel of Figure 4 using pie charts. In the context of an antibody titer of <1:21, only 20% of individuals in the pre-symptomatic group exhibited AIM^+^ responses, while this proportion was notably higher, at 50%, in the pre-inapparent group. Within the antibody titer range of 1:21-1:80, AIM^+^ responses were observed in 33% of pre-symptomatic individuals, contrasting with a substantial 62.5% in the pre-inapparent group. The antibody titers 1:81-1:320 revealed a significant contrast between pre-symptomatic and pre-inapparent donors: while 100% of pre-symptomatic donors exhibited AIM^+^ responses, only 42.85% in the pre-inapparent group did. The inversion in trend can be explained by the small sample size within this bin (n=4) in the pre-symptomatic group, warranting caution in interpreting these findings. Finally, the antibody titer range >1:320 showed notable differences between the percentage of AIM^+^ responders, with 62.50% in the pre-symptomatic group compared to 71.42% in the pre-inapparent group. Importantly, the differences in response frequencies were determined to be statistically significant (p<0.0001) through Fisher’s exact test (**Figure 3**). In conclusion, we found a strong association between the proportion of pre-existing AIM^+^ CD4^+^ T cell memory responders and DENV iELISA titer bins that was more pronounced in the pre-inapparent group than the pre-symptomatic group.

**Figure 3.**
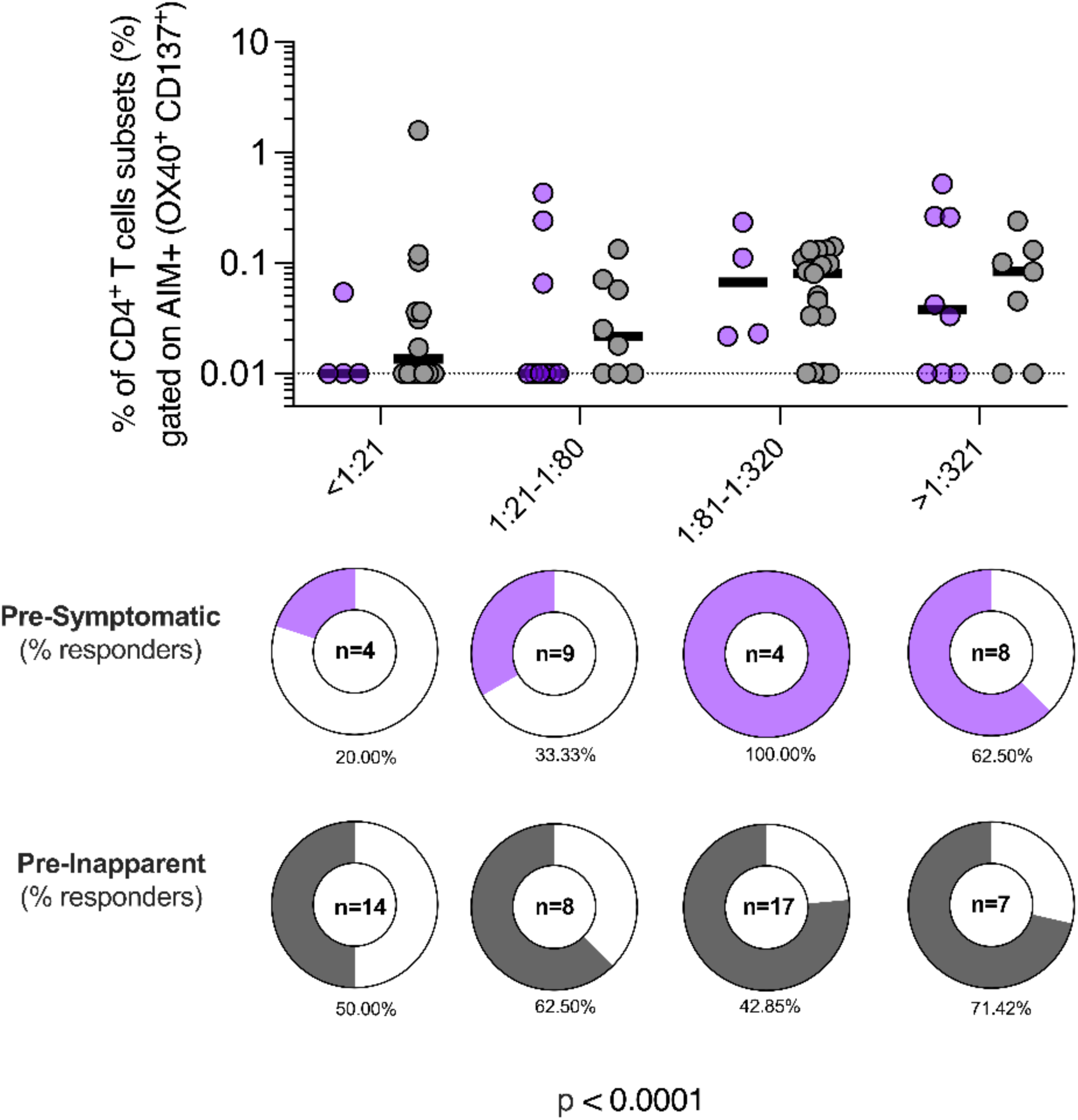
Association of AIM^+^ CD4^+^ T cell responses and pre-existing DENV antibody titers. AIM^+^ CD4^+^ T cell magnitude and pre-existing DENV antibody titer in pre-symptomatic and pre-inapparent groups were plotted side-by-side. AIM^+^ response frequencies are depicted in pie charts. While overall AIM^+^ CD4^+^ T cell responses showed no significant differences (Kruskal-Wallis test) when compared by antibody titer level, statistically significant differences were found when the frequency of responders was compared within the antibody titer bins and between groups using the Fisher’s exact test.

**Figure 4:**
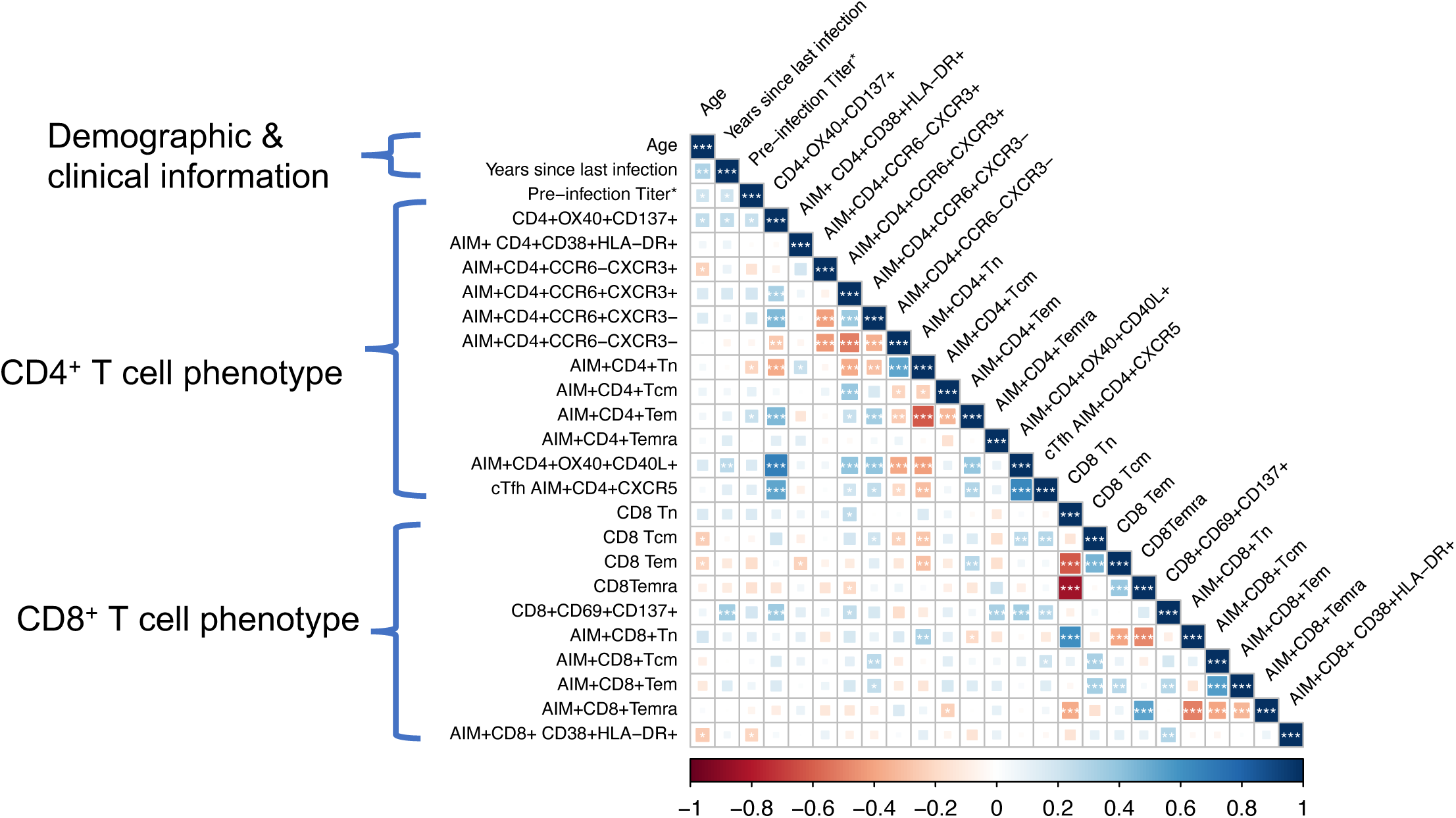
DENV specific T cell immunological correlation analyses. Comprehensive multiparametric analysis of interrelationships among parameters examined to understand immunogenicity from prior DENV infections and its impact on DENV specific immune responses was generated with the corrplot package (version 0.92) in R version 4.2.2. The Spearman correlation method was employed; square size represents absolute correlation coefficient and square color indicates magnitude (red=-1.0, blue=1.0). Significance levels are denoted as *p<0.05, **p<0.01, ***p<0.001, with corresponding p-values indicated by white asterisks.

### HLA associations with disease outcome of the subsequent infection

Polymorphisms within human leukocyte antigen (HLA) alleles, specifically classical HLA-A and HLA-B class I alleles, have been linked to various outcomes in DENV infections, including resistance, susceptibility, and disease severity [23]. Thus, we explored the impact of HLA alleles on the outcome of subsequent DENV infection. Table 2 summarizes results obtained through the application of the RATE tool from the Immune Epitope Database (IEDB). RATE, which stands for Restrictor Analysis Tool for Epitopes, is an automated method designed to infer HLA restriction for a set of given epitopes from large datasets of T cell responses in HLA-typed subjects. The input for this analysis comprised HLA-type information from our donors and the outcome of the subsequent infection. Table 2 shows the allele number and corresponding allele name as denoted in the input data. The columns labeled “A+ Symptomatic+” represent the count of subjects expressing the allele and subsequently experiencing symptomatic infections. The “A+ Symptomatic+ %” column provides the frequency of subjects for each outcome-allele combination within the cohort. Additionally, the “P-value” column displays the P-value obtained from Fisher’s exact test, offering a relative ranking that indicates which HLA restriction has the strongest association with symptomatic outcome. Our findings revealed three notable associations only with subsequent symptomatic outcomes. One of these associations involved HLA-B*51:01, which had previously been associated with the development of dengue hemorrhagic fever (DHF) in patients experiencing secondary infections [21]. Additionally, we also found associations with HLA-A*02:05 and HLA-B*15:10, which had not been previously described in the context of DENV infections. Collectively, our results deliver genetic evidence that underscores the potential role of classical HLA class I alleles in shaping the outcome of subsequent DENV infection (Table 2, Table S2). This table presents associations between HLA-A and HLA-B class I alleles and subsequent DENV infection outcomes. The complete association matrix tables can be found in Table S2.

**Table 2.**
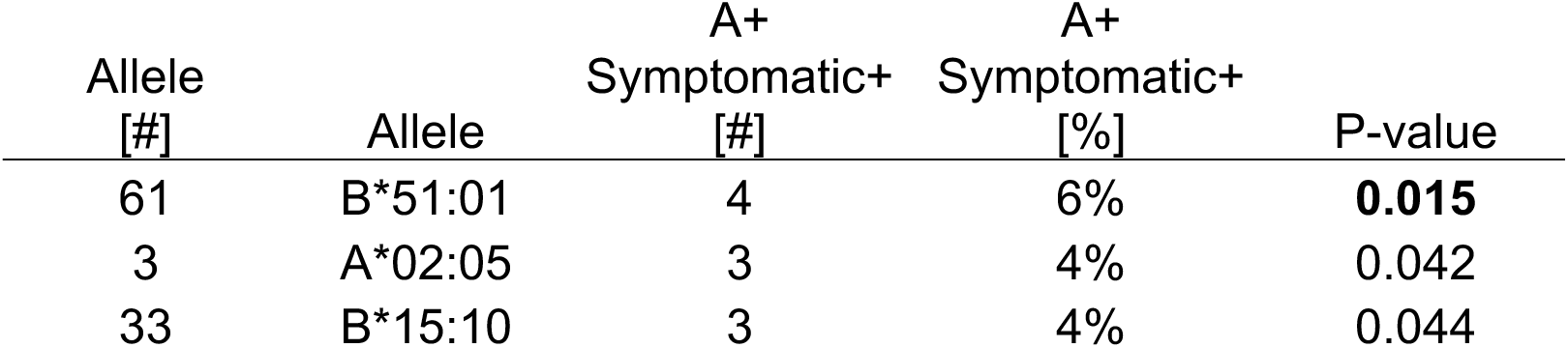
HLA associations with disease outcome of the subsequent infection.

| Allele<br>[#] | Allele | A+<br>Symptomatic+<br>[#] | A+<br>Symptomatic+<br>[%] | P-value |
| --- | --- | --- | --- | --- |
| 61 | B*51:01 | 4 | 6% | **0.015** |
| 3 | A*02:05 | 3 | 4% | 0.042 |
| 33 | B*15:10 | 3 | 4% | 0.044 |

### DENV specific T cell immunological correlation analyses

Finally, we generated a correlation matrix to comprehensively evaluate the interrelationships among the various DENV specific T cell subsets. To ensure an unbiased evaluation of all potential correlations, we included numerical demographic information such as age, years since last infection, pre-infection titer which corresponds to the DENV iELISA, and all parameters used to define T cell populations with our flow cytometry analysis, even if these T cell subsets were rare events that were not strong represented in most of the analyzed donors. This encompassed AIM^+^ CD4^+^ T effector memory cells, recently activated CD4^+^ CD38^+^ HLA-DR^+^ T cells and circulating T follicular helper cells CD4^+^CXCR5^+^ T cells.

The key findings of our analysis confirm a robust positive correlation between the AIM responses CD4^+^OX40^+^CD137^+^ and CD8^+^CD69^+^CD137^+^ T cells. We identified a statistically significant but weaker positive correlation between pre-existing antibody titers and AIM^+^ CD4^+^ T cells. Conversely, no significant association was observed between pre-existing antibody titers and CD8^+^ T cells. Notably, within the CD8^+^ T cell subset, the T_EMRA_ cell subset exhibited a positive and significant correlation with AIM^+^ CD8^+^ T cell responses (CD69^+^CD137^+^).

In summary, our analysis unveiled substantial relationships within T cell memory profiles among individuals naturally infected with DENV. This underscores the distinct T cell memory profiles associated with different outcomes following subsequent DENV infections.

## Discussion

In our study of a long-term pediatric cohort in Managua, Nicaragua, we examined memory T cell responses to naturally acquired DENV infections. We focused on 71 healthy children with one or more prior DENV exposures before a subsequent DENV infection within the following year. Our primary goal was to investigate how pre-existing DENV specific T cell responses impacted the outcome of the following DENV infection: inapparent versus symptomatic. To investigate the presence of DENV specific T cells in these healthy children, we employed the activation-induced marker (AIM) assay. This approach, which is based on T cell receptor (TCR)-dependent activation marker upregulation, allowed us to capture a diverse and cytokine-independent T cell response [22].

Utilizing this method, we detected DENV specific CD4^+^ T cells in 61% and DENV specific CD8^+^ T cells in 59% of all children with prior DENV infection. When considering the entire cohort, we did not identify discernible differences in the levels of overall DENV specific CD4^+^ and CD8^+^ T cell responses between children who subsequently developed symptomatic or inapparent infections. However, when groups were stratified by infection history, DENV specific CD4^+^ and CD8^+^ T cell responses exhibited a higher magnitude after two infections, only in children with subsequent inapparent infections. Furthermore, DENV specific CD8^+^ T cell responses exhibited a higher magnitude after two infections in children with subsequent inapparent infections compared to children with subsequent symptomatic infections. This phenomenon in children with a history of more than two prior DENV infections suggests a potential requirement for multiple DENV infections to induce a robust T cell memory response.

The definitive role of pre-existing DENV specific T cells has not been unanimously defined. While higher pre-existing T cell responses have previously been associated with disease [19, 24, 25], our findings diverge, as we found that higher pre-existing DENV specific T cells were associated with the subsequent inapparent outcome of infection. Notably, the aforementioned study employed a cultured *in vitro* ELISPOT assay that did not distinguish between the relative contributions of different T cell subsets. Additionally, the study cohort was relatively small and the methodology involved the use of peptides with a length suitable to favor CD4^+^ T cell responses. In contrast, in a DENV human challenge model, IFNγ from T cells in acute DENV infection were associated with a protective role in *vivo* [26]. In agreement, our study describes a significant association between the magnitude of DENV specific T cells before a subsequent inapparent DENV infection outcome.

Additionally, the results from our study revealed a significant Th1/Th17 polarization within DENV specific CD4^+^ T cells in children who experienced subsequent symptomatic infections. This subset has previously been described within the memory CD4^+^ T cells from patient cohorts experiencing a latent *Mycobacterium tuberculosis* (Mtb) infection [27] but to the best of our knowledge, it is the first description in the context of DENV T cell memory. In PBMCs from patients experiencing DHF, flow cytometry analysis after nonspecific polyclonal stimulation demonstrated a strong immune response skewed towards IL-17 production, marked by a heightened occurrence of CD4^+^IL-17^+^T cells [28]. This suggests that Th17-polarized cells could contribute to symptomatic disease in DENV infection. Additionally, chemokine receptors could play an important role in mediating T cell recruitment to the site of inflammations. The expression of CCR6 and CXCR3 on T cells reflects their diverse capacities for trafficking to non-lymphoid tissues [29]. The source and differentiation requirements of this particular Th1/Th17 subgroup, along with their potential functions, require further clarification. Future studies should investigate whether this Th1/Th17 subset synergistically collaborates with other pro-inflammatory cytokines, leading to an augmentation of systemic inflammation.

When analyzing the memory cell subsets within the AIM^+^ DENV specific T cells, we found a higher frequency of T effector memory re-expressing CD45RA (T_EMRA_) cells in CD4^+^ T cells and CD8^+^ T cells among children with subsequent inapparent infections. T_EMRA_ cells have been described as a subset of CD4^+^ T cells known for their cytotoxic abilities, as indicated by surface exposure of CD107a, a molecule that is translocated to the outside of the cell membrane upon degranulation and the release of cytotoxic molecules such as granzyme B and perforin [30]. T_EMRA_ T cells have been described by us and others in individuals with both primary and secondary DENV infections, with varying numbers based on infection history. T_EMRA_ CD4^+^ and CD8^+^ T cells have been detected in secondary DENV infections, indicating repeated exposure to DENV antigens triggers their development [24, 31]. Notably, DENV specific T_EMRA_ cells are more abundant in patients with mild dengue fever (DF) compared to severe cases, implying a protective role [16]. In summary, our results demonstrate for the first time that pre-existing CD4^+^ and CD8^+^ T_EMRA_ cells are significantly associated with subsequent symptomatic infection outcome, particularly in those with a history of repeated exposure.

In our study, we describe a significant relationship between HLA types and the subsequent outcome of DENV infections. Our findings not only corroborate the previously reported association involving HLA-B51:01 but also introduce two additional HLA types linked to subsequent symptomatic infections. Understanding HLA-disease associations is crucial, as it sheds light on the genetic factors that influence susceptibility to specific diseases.

In summary, we detected DENV specific CD4^+^ T cells in and DENV specific CD8^+^ T cells in ~60% of children in our study. Notably, children with 2+ prior DENV infections had significantly higher DENV specific CD8^+^ T cells than those with only one prior DENV infection. Furthermore, children with a subsequent symptomatic infection exhibited significant Th1/Th17 polarization in DENV specific CD4^+^ T cells. Additionally, children with inapparent infections had a higher frequency of pre-existing T_EMRA_ T cells in DENV specific CD4^+^ and CD8^+^ T cells. The relationship between higher DENV specific memory T cells and subsequent inapparent DENV infections in children after 2+DENV infections, highlights a protective role for these cells. Importantly, the protective role of pre-existing T cell memory was enhanced in children with multiple prior infections. This is important to consider in the context of DENV vaccination programs, where a number of previous natural infection may occur in endemic settings.

Our study demonstrates the ability to detect antigen-specific T cells in healthy children with a history of DENV infections by analyzing the co-expression patterns of activation-induced markers (AIM) on T cells. Experiencing two or more DENV infections are associated with the presence of DENV specific T cells. Importantly, heightened DENV specific T cell presence was correlated with an absence of clinical symptoms during subsequent DENV infections, suggesting a protective role for DENV specific T cells in DENV infections.

### Limitations of our Study

The study we conducted has several limitations that need to be taken into account when interpreting our findings. The time elapsed since the last DENV infection in our study participants ranged from approximately 3 to 5 years. T cell responses can be highly sensitive to temporal factors and change over time. Our study examined a single time point before a subsequent DENV infection, with two possible outcomes. This means we were not able to describe the dynamic changes in the T cell response over time. Finally, our study focused on examining PBMCs within the bloodstream of healthy children. These cells may not provide an accurate reflection of the DENV specific T cell repertoire developed in response to previous DENV infections, as this repertoire is predominantly localized to tissues like the skin, where the initial virus inoculation occurs. These variations in infection dynamics, initial viral load and the localization of the T cell memory repertoire could potentially influence the outcome of subsequent DENV infections that cannot be assessed with our approach.

## Methods

### Study approval

Samples were collected under the ethical approval provided by the Institutional Review Boards (IRBs) of the La Jolla Institute for Immunology (LJI VD-085), the University of California, Berkeley, and the Nicaraguan Ministry of Health.

### Study population

A total of 103 peripheral blood samples were collected from participants enrolled in the Pediatric Dengue Cohort Study (PDCS), an ongoing prospective cohort spanning two decades, centered on children aged 2 to 17, in Managua, Nicaragua. A description of this cohort has been outlined previously in [32]. To briefly elucidate, upon enrollment, parents committed to promptly seek medical attention for their children upon the emergence of initial symptoms. Validation of cases presenting symptomatic Dengue was accomplished via the identification of DENV RNA using RT-PCR, real-time RT-PCR, and/or virus isolation within the acute-phase sample (within 0-6 days post-symptom onset).

### Serology

Evaluation of inapparent DENV infections is performed annually by comparing serum samples obtained from two consecutive healthy years utilizing the inhibition enzyme-linked immunosorbent assay (iELISA) as detailed in *Katzelnick et al.* [8]. An iELISA titer increase exceeding 4-fold between yearly samples in children who did not seek medical care at the study Health Center between the annual samplings is considered an inapparent infection [33].

### Exclusion criteria

In our study, we initially examined 103 samples, but upon unblinding, 32 donors were identified as “DENV immune”. Due to the uncertainty about the amount of previous DENV infections and the timing of their last infection, we opted to exclude these donors from our analysis. This decision was made to preserve the accuracy of our research, as both the exact number of past DENV infections and the time frame of these infections are pivotal factors in our analysis of the influence of pre-existing T cells in the outcome of subsequent DENV infections.

### PBMC Isolation

Human blood samples were subjected to density gradient centrifugation using Ficoll-Paque Premium (GE Healthcare Biosciences, Kowloon, Hong Kong). Subsequently, the isolated peripheral blood mononuclear cells (PBMCs) were resuspended in fetal bovine serum (Gemini Bio-products, Sacramento, California; Gibco Life Technologies) containing 10% dimethyl sulfoxide and cryopreserved in liquid nitrogen, following established protocols described in [18].

### DENV megapools (DENV MP)

The MegaPool approach was previously established by *Sidney et al.* [34] and recently detailed protocols were made public *R. da Silva Antunes et al.* [17] allowing the simultaneous assessment of numerous T cells from diverse specificity. Briefly, this method consists of the dissolution of a diverse array of epitopes, their consolidation into a combined pool, and subsequent joint lyophilization to avoid cell cytotoxicity by a larger amount of DMSO. In this study, peripheral blood mononuclear cells (PBMCs) were *ex vivo* stimulated to enable the examination of antigen-specific T cell responses against the Dengue Virus using flow cytometry. The employed DENV specific peptide pool for CD8^+^ T cells was composed of 268 overlapping 9 and 10-mers and the DENV specific CD4^+^ T cell peptide pool was composed of 180 overlapping 15-mers as previously described by [19, 35].

### T cell assays

All T cell assays were carried out by two independent investigators (RIG and EAE) on the same Spectral analyzer Cytek Aurora. The samples were distributed in a blinded manner. Samples were run including a positive (PHA) and negative control (DMSO) for each donor in the same experiment. Furthermore, each independent experiment included a sample of the same control donor, with known CD4 and CD8 T cell reactivity. Details of the T cell assays are provided below. After all experiments were run, experimentalists were unblinded to allow for specific analysis and data interpretation.

### Activation-induced marker Assay (AIM)

The AIM assays were performed following established methods described previously by [21]. An overview of the experimental design is provided in supplemental Fig. S1. The quantification of Dengue-specific T cells was determined by assessing the proportion of AIM+ (OX40^+^CD137^+^) CD4^+^ and (CD69^+^CD137^+^) CD8^+^ T cells after a 24h PBMC stimulation. Briefly, before adding the Dengue MP, PBMCs were subjected to a 15 min blocking step at 37°C using 0.5 µg/ml anti-CD40 monoclonal antibody (Miltenyi, Biotec). Following this, cells were incubated with fluorescently labeled chemokine receptor antibodies (anti-CCR6, CXCR5, CXCR3, and CCR7) and the Dengue MPs (1 µg/ml) in 96-well U-bottom plates, and incubated at 37°C, 5 % CO_2_ for 24 hours. Additionally, PBMCs were incubated with an equimolar quantity of DMSO for negative control purposes, and with phytohemagglutinin (PHA, 0.5 µg/ml) (Roche) as a positive control. For surface staining, 1×10^6^ PBMCs were resuspended in PBS, treated with human FC block from BD Bioscience, and stained with LIVE/DEAD Blue marker (Thermo Fisher Scientific) in darkness for 15 minutes, and then rinsed with PBS. Subsequently, the remaining surface antibodies were added to the cells and incubated for 60 minutes at 4°C in the dark. After surface staining, the cells were washed twice with PBS containing 3% FBS (FACS buffer). The cells were immediately acquired using a Cytek Aurora flow cytometer (Cytek Biosciences, Fremont, CA). The antibodies employed in this panel are listed in Table S1, and the representative gating strategy for Dengue-specific CD4^+^ and CD8^+^ T cells using the AIM assay is illustrated in Figure S2.

The quantification of DENV specific CD4^+^ and CD8^+^ T cells was achieved by subtracting the frequency of AIM^+^ cells in the unstimulated condition from the frequency in the antigen-stimulated setting. The minimum DMSO level was fixed at 0.005%. The fold change was calculated as the ratio of the frequency of AIM^+^ cells during antigen stimulation to that in the unstimulated state. Responses above 0.01% and a stimulation index (SI) greater than 1.5 were considered positive.

### HLA typing

HLA typing for Class I (HLA A, B, C) and Class II (DRB1, DRB3/4/5, DQA1/DQB1, DPB1) was carried out by a laboratory accredited by ASHI at Murdoch University in Western Australia, following established protocols as detailed previously, using locus-specific PCR amplification of genomic DNA. Table 1 Lists the HLA types of all donors used in this study.

### RATE analysis

This table presents associations between HLA-A and HLA-B class I alleles and subsequent DENV infection outcomes. The complete association matrix tables can be found in supplemental table S2. The association matrix was generated using the RATE tool, as described in methods. The RATE tool is available online at http://iedb-rate.liai.org The RATE tool was used to compute the HLA restrictions by considering the presence or absence of a given stimulus as the biological outcome. We uploaded the HLA typing information and response data as .txt files. The cohort was divided into pre-inapparent and pre-symptomatic; the individuals classified as symptomatic had positive responses (value = 1), while the inapparent had a negative response (value = 0). The cutoff was established at 1 for the positive response because the response data was provided in binary values. The script calculated the subjects’ relative frequency (RF) to a given stimulus and the expression of a given allele compared to the general test population and associated statistical significance. RF>1 indicates a positive association between the two properties in question, meaning that the expression of the specific allele increases the possibility of having a positive immune response. RATE tool used Fisher’s exact test to estimate the statistical significance of the association between HLA molecules and stimuli responses. The RATE tool is a Python 2.6.5+ CGI script previously described and validated in [36]. The RATE tool is available online at http://iedb-rate.liai.org.

### Correlation Analysis

Correlograms plotting the Spearman rank correlation coefficient between all paired parameters were created with the corrplot package (version 0.92) in R version 4.2.2, as previously described (doi.org/10.1016/j.cell.2022.05.022). Significant correlations were calculated using corr.mtest and graphed based on * p <0.05, ** p <0.01, *** p <0.001.

### Statistical analysis

Flow cytometry data analysis was conducted using FlowJo version 10.8.2. Statistical analyses were carried out using GraphPad Prism version 9.3.0 unless specified otherwise. Specifics of the statistical analyses performed are provided within the corresponding figure description. Data presented in linear scales were represented as Geometric Mean. Unpaired comparisons were evaluated using Mann–Whitney. For multiple comparisons Kruskal–Wallis tests followed by Dunn’s posttest was employed. Additional details regarding significance are included in the respective figure descriptions.

## Materials availability

Requests should be directed to, Dr. Daniela Weiskopf. Upon specific request and execution of a material transfer agreement (MTA), aliquots of the peptide pools utilized in this study can be made available. Limitations might be applied to the availability of peptide reagents due to cost, quantity, demand, and availability.

## Data availability

All the data generated in this study are available in the published article and summarized in the corresponding tables, figures, and supplemental materials.

## Author contributions

Conceptualization: DW, EH; Methodology: RIG; EAE, Formal analysis: RIG, EAE, AMP; Investigation: RIG, EAE, AMP, TS, JVZ, AB, EH, DW; Project administration: EH and DW; Funding acquisition: EH and DW; Writing: RIG, EH, DW; Supervision: DW. All authors provided critical feedback and helped shape the research, analysis, and manuscript.

## Data Availability

All data produced in the present study are available upon reasonable request to the authors
All data produced in the present work are contained in the manuscript

## Acknowledgments

We are very grateful to the Nicaraguan children and their families who contributed samples to the Pediatric Dengue Cohort Study. We would also like to thank the staff at the Sustainable Sciences Institute and the Centro Nacional de Diagnóstico y Referencia, Ministerio de Salud who have been biobanking samples and clinical data for 19 years to make this study on pre-infection samples possible. In addition, we would like to acknowledge Dr. Claudia Sanchez San Martin for her support with sample inventory and shipment, Mr. Raul Zapata for processing these pediatric blood samples, and Mr. Yuri Villabos for his excellence in sample management and inventory.

---

**Figure S1.**
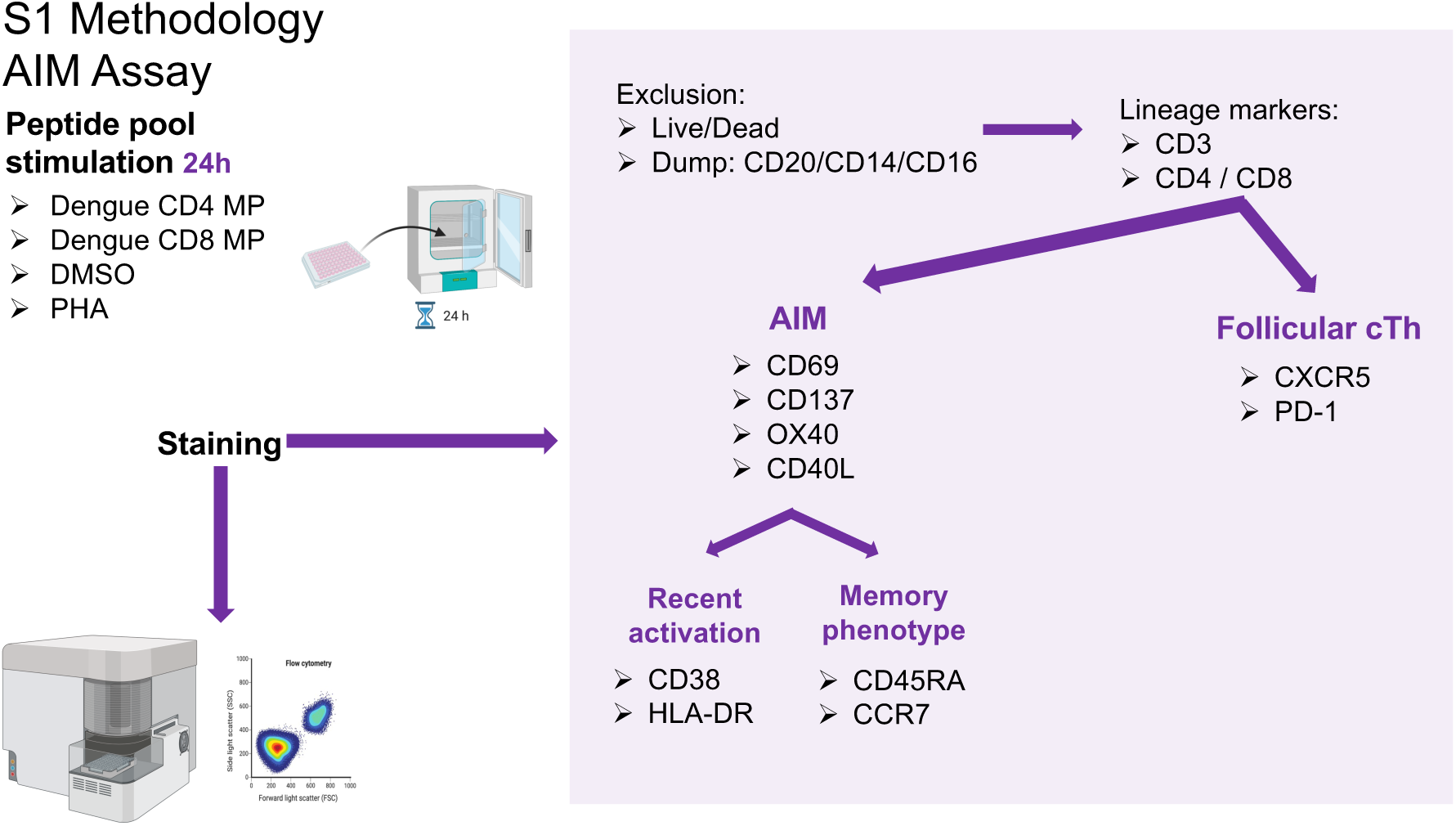
Representative experimental design for AIM Assay for T cell analysis.

**Figure S2.**
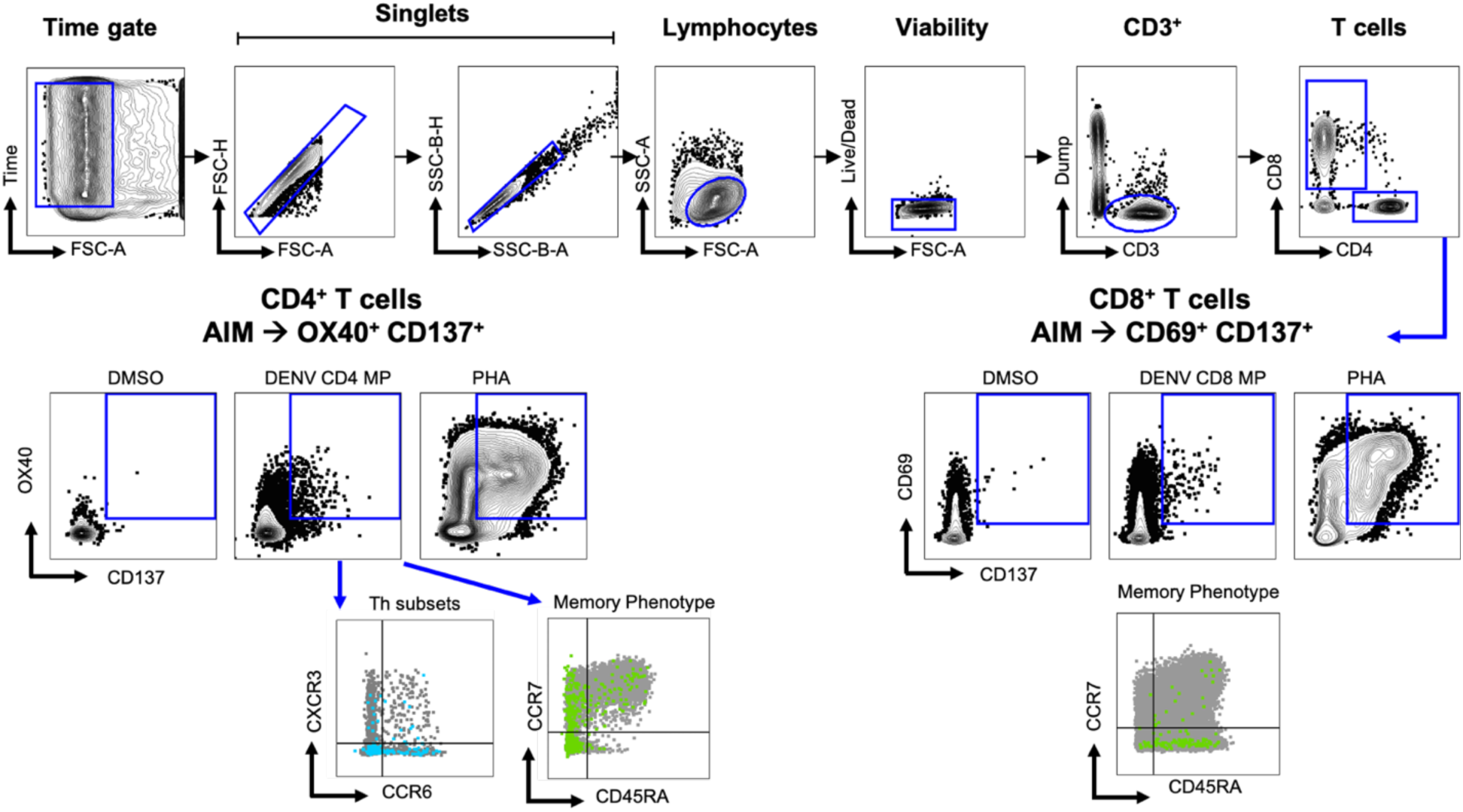
Representative gating strategy for DENV specific T cell analysis. Related to Figures 2–3 (A) Representative strategy to define AIM^+^ CD4^+^ T cells: CD3^+^CD4^+^OX40^+^CD137^+^ (B) Representative strategy to define AIM^+^ CD8^+^ T cells: CD3^+^CD8^+^CD69^+^CD137^+^

## References

1. Reuters, WHO warns of dengue risk as global warming pushes cases near historic highs. 2023.

2. Pierson, T.C. and M.S. Diamond, The continued threat of emerging flaviviruses. Nature Microbiology, 2020. 5(6): p. 796–812.

3. Sharp, T.M., et al., Knowledge gaps in the epidemiology of severe dengue impede vaccine evaluation. The Lancet Infectious Diseases, 2022. 22(2): p. e42–e51.

4. St. John, A.L. and A.P.S. Rathore, Adaptive immune responses to primary and secondary dengue virus infections. Nature Reviews Immunology, 2019. 19(4): p. 218–230.

5. Teo, A., et al., Understanding antibody-dependent enhancement in dengue: Are afucosylated IgG1s a concern? PLOS Pathogens, 2023. 19(3): p. e1011223.

6. Katzelnick, L.C., et al., Neutralizing antibody titers against dengue virus correlate with protection from symptomatic infection in a longitudinal cohort. Proceedings of the National Academy of Sciences, 2016. 113(3): p. 728–733.

7. Katzelnick, L.C., S. Bos, and E. Harris, Protective and enhancing interactions among dengue viruses 1-4 and Zika virus. Current Opinion in Virology, 2020. 43: p. 59–70.

8. Katzelnick, L.C., et al., Antibody-dependent enhancement of severe dengue disease in humans. Science, 2017. 358(6365): p. 929–932.

9. Salje, H., et al., Reconstruction of antibody dynamics and infection histories to evaluate dengue risk. Nature, 2018. 557(7707): p. 719–723.

10. Weiskopf, D., et al., Comprehensive analysis of dengue virus-specific responses supports an HLA-linked protective role for CD8+ T cells. Proceedings of the National Academy of Sciences, 2013. 110(22): p. E2046–E2053.

11. Rivino, L., et al., Virus-specific T lymphocytes home to the skin during natural dengue infection. Science Translational Medicine, 2015. 7(278): p. 278ra35–278ra35.

12. Künzli, M. and D. Masopust, CD4+ T cell memory. Nature Immunology, 2023. 24(6): p. 903–914.

13. Rothman, A.L., Immunity to dengue virus: a tale of original antigenic sin and tropical cytokine storms. Nature Reviews Immunology, 2011. 11(8): p. 532–543.

14. Farber, D.L., N.A. Yudanin, and N.P. Restifo, Human memory T cells: generation, compartmentalization and homeostasis. Nat Rev Immunol, 2014. 14(1): p. 24–35.

15. Mongkolsapaya, J., et al., Original antigenic sin and apoptosis in the pathogenesis of dengue hemorrhagic fever. Nature Medicine, 2003. 9(7): p. 921–927.

16. Duangchinda, T., et al., Immunodominant T-cell responses to dengue virus NS3 are associated with DHF. Proceedings of the National Academy of Sciences, 2010. 107(39): p. 16922–16927.

17. da Silva Antunes, R., et al., The MegaPool Approach to Characterize Adaptive CD4+ and CD8+ T Cell Responses. Current Protocols, 2023. 3(11): p. e934.

18. Weiskopf, D., et al., Human CD8+ T-Cell Responses Against the 4 Dengue Virus Serotypes Are Associated With Distinct Patterns of Protein Targets. The Journal of Infectious Diseases, 2015. 212(11): p. 1743–1751.

19. Grifoni, A., et al., Global Assessment of Dengue Virus-Specific CD4+ T Cell Responses in Dengue-Endemic Areas. Frontiers in Immunology, 2017. 8.

20. Zhang, Z., et al., Humoral and cellular immune memory to four COVID-19 vaccines. Cell, 2022.

21. Reiss, S., et al., Comparative analysis of activation induced marker (AIM) assays for sensitive identification of antigen-specific CD4 T cells. PLOS ONE, 2017. 12(10): p. e0186998.

22. Poloni, C., et al., T-cell activation–induced marker assays in health and disease. Immunology & Cell Biology, 2023. 101(6): p. 491–503.

23. Dendrou, C.A., et al., HLA variation and disease. Nature Reviews Immunology, 2018. 18(5): p. 325–339.

24. Tian, Y., et al., Dengue-specific CD8+ T cell subsets display specialized transcriptomic and TCR profiles. The Journal of Clinical Investigation, 2019. 129(4): p. 1727–1741.

25. Grifoni, A., et al., Transcriptomics of Acute DENV-Specific CD8+ T Cells Does Not Support Qualitative Differences as Drivers of Disease Severity. Vaccines, 2022. 10(4): p. 612.

26. Gunther, V.J., et al., A human challenge model for dengue infection reveals a possible protective role for sustained interferon gamma levels during the acute phase of illness. Vaccine, 2011. 29(22): p. 3895–3904.

27. Lindestam Arlehamn, C.S., et al., Memory T Cells in Latent Mycobacterium tuberculosis Infection Are Directed against Three Antigenic Islands and Largely Contained in a CXCR3+CCR6+ Th1 Subset. PLOS Pathogens, 2013. 9(1): p. e1003130.

28. Sánchez-Vargas, L.A., et al., Characterization of the IL-17 and CD4+ Th17 Cells in the Clinical Course of Dengue Virus Infections. Viruses, 2020. 12(12): p. 1435.

29. Becattini, S., et al., Functional heterogeneity of human memory CD4^+^ T cell clones primed by pathogens or vaccines. Science, 2015. 347(6220): p. 400–406.

30. Weiskopf, D., et al., Dengue virus infection elicits highly polarized CX3CR1+ cytotoxic CD4+ T cells associated with protective immunity. Proceedings of the National Academy of Sciences, 2015. 112(31): p. E4256–E4263.

31. Tian, Y., et al., Molecular Signatures of Dengue Virus-Specific IL-10/IFN-γ Co-producing CD4 T Cells and Their Association with Dengue Disease. Cell Reports, 2019. 29(13): p. 4482–4495.e4.

32. Kuan, G., et al., The Nicaraguan Pediatric Dengue Cohort Study: Study Design, Methods, Use of Information Technology, and Extension to Other Infectious Diseases. American Journal of Epidemiology, 2009. 170(1): p. 120–129.

33. Gordon, A., et al., The Nicaraguan Pediatric Dengue Cohort Study: Incidence of Inapparent and Symptomatic Dengue Virus Infections, 2004–2010. PLOS Neglected Tropical Diseases, 2013. 7(9): p. e2462.

34. Sidney, J., B. Peters, and A. Sette, Epitope prediction and identification-adaptive T cell responses in humans. Semin Immunol, 2020. 50: p. 101418.

35. Weiskopf, D., et al., Comprehensive analysis of dengue virus-specific responses supports an HLA-linked protective role for CD8+ T cells. Proceedings of the National Academy of Sciences, 2013. 110(22): p. E2046–E2053.

36. Paul, S., et al., A Population Response Analysis Approach To Assign Class II HLA-Epitope Restrictions. The Journal of Immunology, 2015. 194(12): p. 6164–6176.

