## Supplementary material for "Frequency of Dengue Virus-Specific T Cells is related to Infection Outcome in Endemic Settings": Suplemental Table S1

### Table S1. Antibodies used for AIM assays

| Marker | Fluorochrome | Clone | Source | Catalog. No. | Dilution |
|---|---|---|---|---|---|
| Live/Dead | Blue | | ThermoFisher | L23105 | 1:1000 |
| CCR6 | BUV496 | 11A9 | BD Biosciences | 612948 | 1:200 |
| CCR7 | BV711 | G043H7 | Biolegend | 353228 | 1:200 |
| CD137 | BUV737 | 4B4-1 | BD Biosciences | 741861 | 1:100 |
| CD14 | BV510 | 63D3 | Biolegend | 367124 | 1:1000 |
| CD16 | BV510 | 3G8 | Biolegend | 302048 | 1:1000 |
| CD20 | BV510 | 2H7 | Biolegend | 302340 | 1:1000 |
| CD3- | BUV395 | UCHT1 | BD Biosciences | 563546 | 1:1000 |
| CD38 | BV650 | HB-7 | Biolegend | 356620 | 1:200 |
| CD4 | cFluor b548 | SK3 | Cytek Biosciences | R7-20043 | 1:500 |
| CD40L | PE-Dazzle594 | 24-31 | Biolegend | 310840 | 1:200 |
| CD45RA | BV570 | HI100 | Biolegend | 304132 | 1:1000 |
| CD69 | FITC | FN50 | Biolegend | 310904 | 1:200 |
| CD8 | BUV805 | SK1 | BD Biosciences | 612889 | 1:1000 |
| CD95 | BB700 | DX2 | BD Biosciences | 566542 | 1:500 |
| CXCR3 | BV605 | G025H7 | Biolegend | 353728 | 1:200 |
| CXCR5 | BV421 | J252D4 | Biolegend | 356920 | 1:200 |
| HLA-DR | APC-R700 | G46-6 | BD Biosciences | 565127 | 1:500 |
| OX40 | APC | Ber-Act35 | Biolegend | 350008 | 1:100 |
| PD-1 | BV785 | EH12.2H7 | Biolegend | 329930 | 1:200 |
