## Supplementary material for "Frequency of Dengue Virus-Specific T Cells is related to Infection Outcome in Endemic Settings": Suplemental Table S2

**Table S2 HLA Phenotype and RATE Tool results for each donor.**

| Allele | A+R+ | A+R+ % | No_of_Donors | Relative_freq | P-value |
|---|---|---|---|---|---|
| **A*01:01** | 3 | 0.04 | 70 | 1.680 | 0.341 |
| **A*02:01** | 10 | 0.14 | 70 | 1.000 | 1.000 |
| **A*02:05** | 3 | 0.04 | 70 | 2.800 | 0.042 |
| **A*02:06** | 1 | 0.01 | 70 | 0.700 | 1.000 |
| **A*02:33** | 0 | 0 | 70 | 0.000 | 1.000 |
| **A*03:01** | 3 | 0.04 | 70 | 0.933 | 1.000 |
| **A*11:01** | 0 | 0 | 70 | 0.000 | 0.534 |
| **A*23:01** | 1 | 0.01 | 70 | 0.700 | 1.000 |
| **A*23:05** | 0 | 0 | 70 | 0.000 | 1.000 |
| **A*24:02** | 8 | 0.11 | 70 | 0.896 | 0.795 |
| **A*25:01** | 0 | 0 | 70 | 0.000 | 0.534 |
| **A*26:01** | 1 | 0.01 | 70 | 0.933 | 1.000 |
| **A*30:01** | 1 | 0.01 | 70 | 2.800 | 0.357 |
| **A*30:02** | 1 | 0.01 | 70 | 0.467 | 0.410 |
| **A*31:01** | 2 | 0.03 | 70 | 1.120 | 1.000 |
| **A*32:01** | 1 | 0.01 | 70 | 2.800 | 0.357 |
| **A*33:01** | 1 | 0.01 | 70 | 0.933 | 1.000 |
| **A*33:03** | 2 | 0.03 | 70 | 2.800 | 0.124 |
| **A*34:02** | 0 | 0 | 70 | 0.000 | 1.000 |
| **A*68:01** | 3 | 0.04 | 70 | 1.050 | 1.000 |
| **A*68:02** | 2 | 0.03 | 70 | 1.400 | 0.613 |
| **A*68:03** | 3 | 0.04 | 70 | 1.200 | 0.694 |
| **A*68:07** | 0 | 0 | 70 | 0.000 | 1.000 |
| **A*74:01** | 0 | 0 | 70 | 0.000 | 1.000 |
| **B*07:02** | 1 | 0.01 | 69 | 0.460 | 0.406 |
| **B*08:01** | 1 | 0.01 | 69 | 0.690 | 1.000 |
| **B*13:02** | 1 | 0.01 | 69 | 2.760 | 0.362 |
| **B*14:01** | 0 | 0 | 69 | 0.000 | 1.000 |
| **B*14:02** | 1 | 0.01 | 69 | 0.552 | 0.646 |
| **B*15:01** | 0 | 0 | 69 | 0.000 | 0.289 |
| **B*15:03** | 0 | 0 | 69 | 0.000 | 0.531 |
| **B*15:08** | 0 | 0 | 69 | 0.000 | 1.000 |
| **B*15:10** | 3 | 0.04 | 69 | 2.760 | 0.044 |
| **B*15:15** | 2 | 0.03 | 69 | 1.380 | 0.617 |
| **B*15:30** | 0 | 0 | 69 | 0.000 | 1.000 |
| **B*18:01** | 0 | 0 | 69 | 0.000 | 1.000 |
| **B*27:05** | 0 | 0 | 69 | 0.000 | 1.000 |
| **B*35:01** | 2 | 0.03 | 69 | 0.690 | 0.701 |
| **B*35:12** | 1 | 0.01 | 69 | 0.552 | 0.646 |
| **B*35:17** | 1 | 0.01 | 69 | 0.920 | 1.000 |
| **B*35:20** | 1 | 0.01 | 69 | 2.760 | 0.362 |
| **B*35:23** | 0 | 0 | 69 | 0.000 | 1.000 |
| **B*35:26** | 1 | 0.01 | 69 | 1.380 | 1.000 |

| | | | | | |
|---|---|---|---|---|---|
| **B*35:43** | 3 | 0.04 | 69 | 1.183 | 0.698 |
| **B*38:01** | 1 | 0.01 | 69 | 0.920 | 1.000 |
| **B*39:02** | 1 | 0.01 | 69 | 0.920 | 1.000 |
| **B*39:05** | 0 | 0 | 69 | 0.000 | 0.531 |
| **B*39:08** | 0 | 0 | 69 | 0.000 | 0.289 |
| **B*39:11** | 1 | 0.01 | 69 | 1.380 | 1.000 |
| **B*40:01** | 0 | 0 | 69 | 0.000 | 1.000 |
| **B*40:02** | 7 | 0.1 | 69 | 1.288 | 0.375 |
| **B*40:04** | 0 | 0 | 69 | 0.000 | 1.000 |
| **B*40:11** | 1 | 0.01 | 69 | 1.380 | 1.000 |
| **B*41:01** | 0 | 0 | 69 | 0.000 | 1.000 |
| **B*41:02** | 1 | 0.01 | 69 | 2.760 | 0.362 |
| **B*44:02** | 0 | 0 | 69 | 0.000 | 1.000 |
| **B*44:03** | 1 | 0.01 | 69 | 0.552 | 0.646 |
| **B*45:01** | 0 | 0 | 69 | 0.000 | 1.000 |
| **B*49:01** | 2 | 0.03 | 69 | 1.840 | 0.296 |
| **B*50:01** | 2 | 0.03 | 69 | 1.840 | 0.296 |
| **B*51:01** | 4 | 0.06 | 69 | 2.760 | 0.015 |
| **B*52:01** | 2 | 0.03 | 69 | 0.789 | 1.000 |
| **B*53:01** | 2 | 0.03 | 69 | 2.760 | 0.128 |
| **B*56:01** | 0 | 0 | 69 | 0.000 | 1.000 |
| **B*57:01** | 0 | 0 | 69 | 0.000 | 1.000 |
| **B*58:01** | 0 | 0 | 69 | 0.000 | 0.531 |
| **C*01:02** | 4 | 0.06 | 70 | 1.018 | 1.000 |
| **C*02:02** | 1 | 0.01 | 70 | 0.700 | 1.000 |
| **C*02:10** | 0 | 0 | 70 | 0.000 | 0.534 |
| **C*03:02** | 0 | 0 | 70 | 0.000 | 1.000 |
| **C*03:03** | 3 | 0.04 | 70 | 0.764 | 0.735 |
| **C*03:04** | 6 | 0.09 | 70 | 1.400 | 0.325 |
| **C*03:05** | 4 | 0.06 | 70 | 1.400 | 0.443 |
| **C*04:01** | 8 | 0.11 | 70 | 1.018 | 1.000 |
| **C*05:01** | 0 | 0 | 70 | 0.000 | 1.000 |
| **C*06:02** | 4 | 0.06 | 70 | 1.867 | 0.177 |
| **C*07:01** | 4 | 0.06 | 70 | 1.400 | 0.443 |
| **C*07:02** | 2 | 0.03 | 70 | 0.373 | 0.066 |
| **C*07:05** | 0 | 0 | 70 | 0.000 | 1.000 |
| **C*07:41** | 0 | 0 | 70 | 0.000 | 1.000 |
| **C*08:02** | 1 | 0.01 | 70 | 0.467 | 0.410 |
| **C*12:03** | 2 | 0.03 | 70 | 1.120 | 1.000 |
| **C*15:02** | 1 | 0.01 | 70 | 2.800 | 0.357 |
| **C*16:01** | 3 | 0.04 | 70 | 1.680 | 0.341 |
| **C*17:01** | 1 | 0.01 | 70 | 2.800 | 0.357 |
| **DPB1*01:01** | 3 | 0.04 | 70 | 1.200 | 0.694 |
| **DPB1*02:01** | 3 | 0.04 | 70 | 0.840 | 1.000 |
| **DPB1*03:01** | 3 | 0.04 | 70 | 0.840 | 1.000 |

| | | | | | |
|---|---|---|---|---|---|
| **DPB1*04:01** | 4 | 0.06 | 70 | 0.700 | 0.383 |
| **DPB1*04:02** | 15 | 0.21 | 70 | 0.933 | 0.611 |
| **DPB1*05:01** | 1 | 0.01 | 70 | 0.700 | 1.000 |
| **DPB1*06:01** | 1 | 0.01 | 70 | 2.800 | 0.357 |
| **DPB1*10:01** | 1 | 0.01 | 70 | 1.400 | 1.000 |
| **DPB1*13:01** | 1 | 0.01 | 70 | 0.700 | 1.000 |
| **DPB1*14:01** | 4 | 0.06 | 70 | 1.244 | 0.712 |
| **DPB1*17:01** | 2 | 0.03 | 70 | 1.120 | 1.000 |
| **DPB1*18:01** | 0 | 0 | 70 | 0.000 | 1.000 |
| **DPB1*85:01** | 1 | 0.01 | 70 | 2.800 | 0.357 |
| **DQA1*01:01** | 3 | 0.04 | 70 | 0.840 | 1.000 |
| **DQA1*01:02** | 4 | 0.06 | 70 | 1.018 | 1.000 |
| **DQA1*01:03** | 0 | 0 | 70 | 0.000 | 0.534 |
| **DQA1*02:01** | 3 | 0.04 | 70 | 2.100 | 0.127 |
| **DQA1*03:01** | 21 | 0.3 | 70 | 1.176 | 0.103 |
| **DQA1*04:01** | 2 | 0.03 | 70 | 0.431 | 0.116 |
| **DQA1*05:01** | 8 | 0.11 | 70 | 0.896 | 0.795 |
| **DQB1*02:01** | 1 | 0.01 | 69 | 0.690 | 1.000 |
| **DQB1*02:02** | 4 | 0.06 | 69 | 1.840 | 0.180 |
| **DQB1*03:01** | 8 | 0.12 | 69 | 1.227 | 0.409 |
| **DQB1*03:02** | 17 | 0.25 | 69 | 1.043 | 0.796 |
| **DQB1*04:02** | 4 | 0.06 | 69 | 0.649 | 0.256 |
| **DQB1*05:01** | 4 | 0.06 | 69 | 0.849 | 0.756 |
| **DQB1*05:02** | 1 | 0.01 | 69 | 1.380 | 1.000 |
| **DQB1*05:03** | 0 | 0 | 69 | 0.000 | 1.000 |
| **DQB1*06:02** | 2 | 0.03 | 69 | 0.789 | 1.000 |
| **DQB1*06:03** | 0 | 0 | 69 | 0.000 | 1.000 |
| **DQB1*06:09** | 0 | 0 | 69 | 0.000 | 0.531 |
| **DRB1*01:01** | 1 | 0.01 | 70 | 0.933 | 1.000 |
| **DRB1*01:02** | 1 | 0.01 | 70 | 1.400 | 1.000 |
| **DRB1*01:03** | 0 | 0 | 70 | 0.000 | 1.000 |
| **DRB1*03:01** | 1 | 0.01 | 70 | 0.700 | 1.000 |
| **DRB1*03:02** | 1 | 0.01 | 70 | 0.933 | 1.000 |
| **DRB1*04:02** | 1 | 0.01 | 70 | 0.933 | 1.000 |
| **DRB1*04:03** | 2 | 0.03 | 70 | 1.120 | 1.000 |
| **DRB1*04:04** | 2 | 0.03 | 70 | 1.120 | 1.000 |
| **DRB1*04:05** | 0 | 0 | 70 | 0.000 | 1.000 |
| **DRB1*04:07** | 13 | 0.19 | 70 | 1.174 | 0.452 |
| **DRB1*04:10** | 1 | 0.01 | 70 | 1.400 | 1.000 |
| **DRB1*04:11** | 2 | 0.03 | 70 | 1.120 | 1.000 |
| **DRB1*04:17** | 0 | 0 | 70 | 0.000 | 1.000 |
| **DRB1*07:01** | 4 | 0.06 | 70 | 2.240 | 0.051 |
| **DRB1*08:02** | 1 | 0.01 | 70 | 0.255 | 0.083 |
| **DRB1*08:07** | 0 | 0 | 70 | 0.000 | 1.000 |
| **DRB1*09:01** | 1 | 0.01 | 70 | 1.400 | 1.000 |

| | | | | | |
|---|---|---|---|---|---|
| **DRB1*10:01** | 0 | 0 | 70 | 0.000 | 0.534 |
| **DRB1*11:01** | 4 | 0.06 | 70 | 1.867 | 0.177 |
| **DRB1*11:02** | 1 | 0.01 | 70 | 2.800 | 0.357 |
| **DRB1*11:04** | 2 | 0.03 | 70 | 2.800 | 0.124 |
| **DRB1*12:01** | 0 | 0 | 70 | 0.000 | 0.289 |
| **DRB1*13:01** | 0 | 0 | 70 | 0.000 | 0.534 |
| **DRB1*13:02** | 0 | 0 | 70 | 0.000 | 0.289 |
| **DRB1*13:03** | 1 | 0.01 | 70 | 2.800 | 0.357 |
| **DRB1*13:04** | 2 | 0.03 | 70 | 1.867 | 0.289 |
| **DRB1*14:01** | 0 | 0 | 70 | 0.000 | 1.000 |
| **DRB1*14:02** | 2 | 0.03 | 70 | 1.400 | 0.613 |
| **DRB1*14:06** | 0 | 0 | 70 | 0.000 | 0.082 |
| **DRB1*15:01** | 1 | 0.01 | 70 | 0.700 | 1.000 |
| **DRB1*15:03** | 0 | 0 | 70 | 0.000 | 0.534 |
| **DRB1*16:02** | 1 | 0.01 | 70 | 2.800 | 0.357 |
| **DRB3*01:01** | 4 | 0.06 | 67 | 0.766 | 0.544 |
| **DRB3*02:02** | 9 | 0.13 | 67 | 1.269 | 0.401 |
| **DRB3*03:01** | 0 | 0 | 67 | 0.000 | 0.289 |
| **DRB4*01:01** | 21 | 0.31 | 67 | 1.082 | 0.381 |
| **DRB4*01:03** | 0 | 0 | 67 | 0.000 | 0.525 |
| **DRB5*01:01** | 1 | 0.01 | 67 | 0.536 | 0.643 |
| **DRB5*02:02** | 1 | 0.01 | 67 | 2.680 | 0.373 |
